# Hazard-aware adaptations bridge the generalization gap in large language models: a nationwide study

**DOI:** 10.1101/2025.02.14.25322312

**Authors:** Julie Wu, Sydney Conover, Chloe Su, June Corrigan, John Culnan, Yuhan Liu, Michael Kelley, Nhan Do, Shipra Arya, Alex Sox-Harris, Curtis Langlotz, Renda Weiner, Westyn Branch-Elliman, Summer Han, Nathanael Fillmore

## Abstract

Despite growing excitement in deploying large language models (LLMs) for healthcare, most machine learning studies show success on the same few limited public data sources. It is unclear if and how most results generalize to real-world clinical settings. To measure this gap and shorten it, we analyzed protected notes from over 100 Veterans Affairs (VA) sites, focusing on extracting smoking history—a persistent and clinically impactful problem in natural language processing (NLP). Here we applied adaptation techniques to an LLM over two institutional datasets, a popular public dataset (MIMIC-III) and our VA one, across five smoking history NLP tasks of varying complexity. We demonstrate that adapted prompts, engineered to address observed errors, achieve better generalizability across institutions compared to zero-shot prompts. We analyzed 2,955 notes and LLM outputs to codify errors in a hazard framework, identifying whether error frequency differences between institutions stemmed from generalization failures or inherent data differences. While overall accuracy with the adapted prompt was similar between institutions (macro-F1=0.86 in VA, 0.85 in MIMIC), hazard distributions varied significantly. In some cases, a dataset had more errors in a specific category due to a higher prevalence of the associated hazard, such as templated information in VA notes (p_adj_=0.004). However, when task-specific requirements conflicted with pre-trained model behavior, errors in the untrained institution were more frequent despite similar hazard prevalence (p_adj_=0.007), showing a limit of LLM generalizability. As a potential clinical application, our adapted LLM system identified lung cancer screening eligibility in 59% of Veterans who later developed the disease, compared to 8% with current national VA tools. Our results demonstrate LLM generalizability on real-world, national patient data while identifying hazards to address for improved performance and broader applicability.

## Introduction

With increasing interest in the use of large language models (LLMs) in healthcare settings, from drafting documentation^1,2^ to aiding diagnosis^3,4^, a high bar for evaluation and validation of these LLMs is required. Recent reviews have highlighted the need to use real patient data for evaluation, rather than generated data sources such as medical examination questions^5,6^. However, in a systemic review of 519 studies, Bedi et al. found that the use of real patient data was shockingly low at 5% of these studies due to privacy and ethical concerns^5^. Of the studies that use real patient data, most rely on a small number of public datasets, which are predominantly sourced within single contexts such as intensive care unit stays^7^. Thus, there exists a major barrier in proper evaluation of LLMs using real patient data across diverse disease and hospital settings.

The lack of evaluation on real patient data casts doubt on the relevance of current LLM results to clinical practice. Clinical machine learning models often underperform on new data sources, with some arguing universal generalizability—the ability to transfer performance from training to new settings—is unattainable^8^. Prior algorithms have struggled with issues like performance degradation over time^9^ and limited portability across institutions^10,11^. LLMs, trained on diverse and extensive datasets^12^, offer a potential solution to these challenges, with demonstrated proficiency in natural language processing (NLP)^6,12–14^. NLP is especially beneficial in extracting valuable information embedded in the unstructured clinical notes that can be inaccurate or absent in structured data^15,16^. However, the reliance of many LLMs on generated data or limited healthcare data leaves their ability generalize to real protected healthcare settings unknown.

To bridge the gap between public and protected data, we compared clinical notes from the MIMIC-III database, the most popular publicly available clinical dataset^7,17^, and protected data from the Veteran’s Health Administration (VA), a national network of over 100 hospital systems and 1,000 facilities^18^. We selected smoking history NLP extraction for its clinical significance and technical range. Methodologically, it includes multiple tasks like smoking status, duration, and pack-years, ranging in complexity from straightforward named entity extraction to advanced temporal reasoning^19^. Clinically, it determines lung cancer screening eligibility, which can reduce mortality from the leading cause of cancer-related deaths in the U.S.^20^. However, fewer than 10% of eligible individuals are screened, with the lack of automated identification of smoking history cited as a major barrier^21^. Traditional smoking NLP algorithms, often trained on public or single-institution datasets^22–27^, have faced challenges in generalizing across diverse hospital systems and remain unsolved within the VA^11,22,28^.

To explore the extent and limits to LLM generalization on healthcare data, we tested a range of tasks for an indicative example, smoking history. We go beyond measuring accuracy to illustrate differential hazards unique to real, protected VA patient data that could lead to errors in LLM evaluation versus using publicly available datasets, as well as its potential impact on LLM performance. Finally, we apply prompt engineering to address hazards to LLM generalizability, creating opportunities for NLP-driven clinical applications to improve the identification of lung cancer screening eligibility and facilitate nationwide outreach.

## Results

### Defining tasks and datasets for testing generalizability

To evaluate the generalizability of LLMs for NLP data extraction, we designed smoking history annotation guidelines for five tasks, ranging in complexity and anticipated challenges (Table 1). Included was advanced temporal reasoning, requiring the system to extract all aspects of smoking intervals and their relationships directly from raw text (“end-to-end”), a challenge for prior NLP efforts^29^.

**Table 1.**
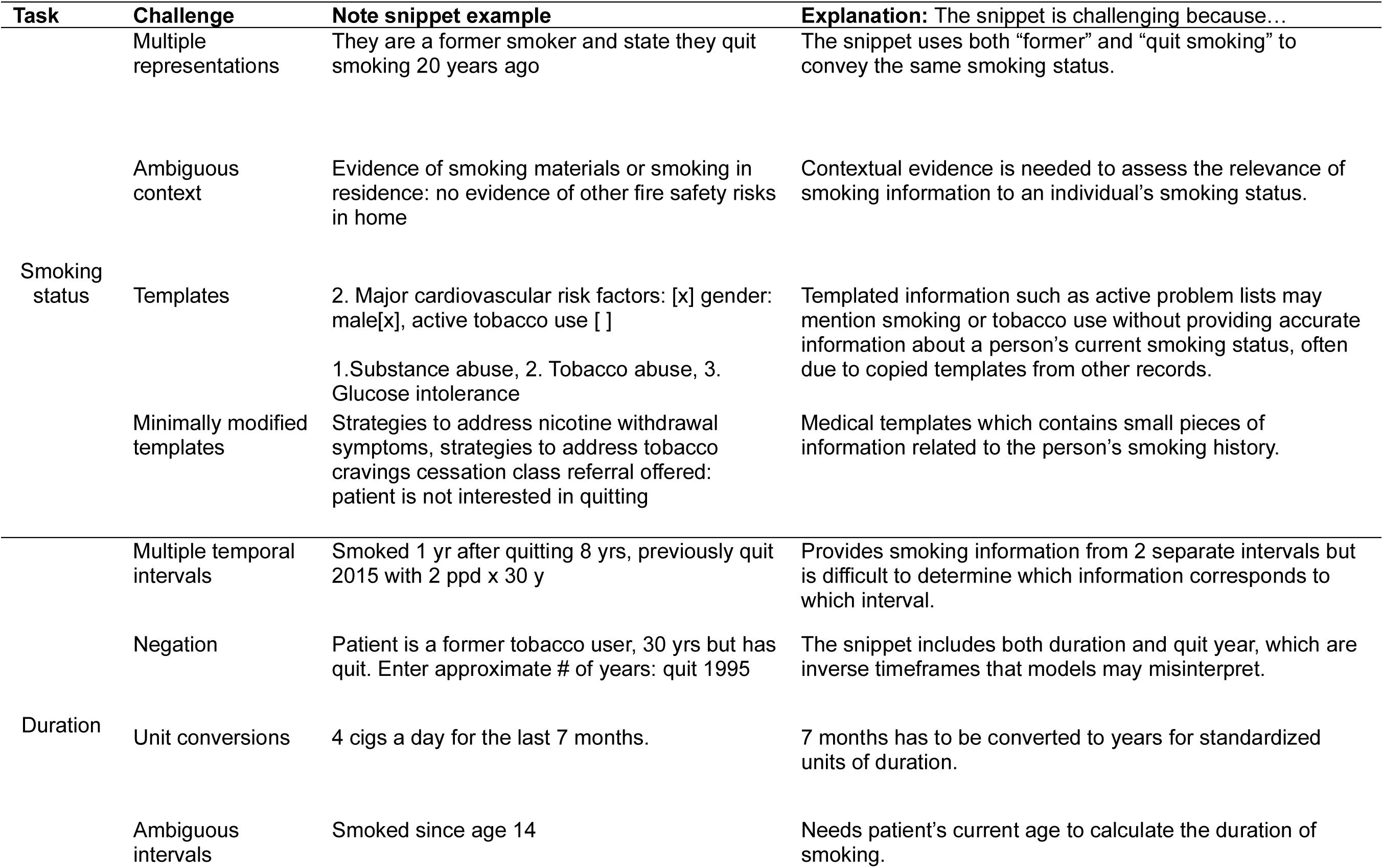
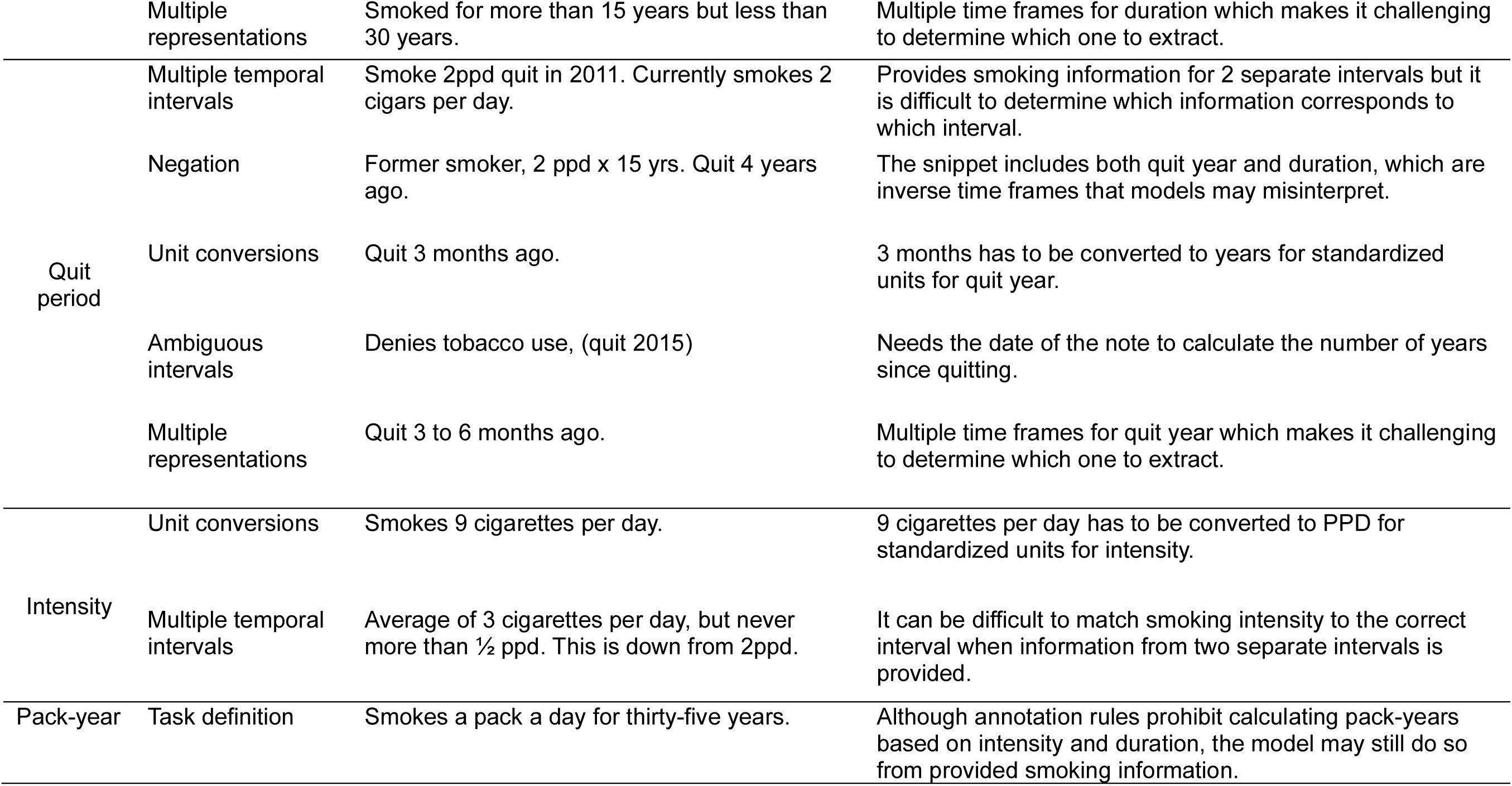
Description of task complexity with example anticipated challenges from clinical notes.

To evaluate how LLMs may perform in a realistic clinical setting, we used clinical notes from the national VA healthcare system. We evaluated the performance of the open-access LLM Mixtral 8×22B, selected for its leading performance-to-cost ratio at the time of the study (April 2024) and availability within the protected VA environment^30,31^.

Notably, VA clinical note data has not been included in any publicly available datasets to date. As a comparative dataset, we included MIMIC-III, a de-identified dataset from a single non-VA institution commonly used in smoking NLP studies^7,17,25,27^.

### Performance of zero-shot prompting varies by task and across institutions

We first tested the generalizability of a zero-shot prompt, modeled after designs that demonstrated success on other tasks and datasets^32,33^, for our study. Task generalizability was defined as the absolute F1 score difference between smoking tasks, indicating robustness to task-specific differences, while institution generalizability was defined as the absolute F1 score difference between the VA (primary validation) and MIMIC (external validation) datasets, indicating robustness to dataset-specific differences. Performance was assessed on 500 note snippets from each cohort. Table 2 summarizes label prevalence for the five smoking history tasks across datasets. Label distributions were similar, except for smoking status, with more current smokers and fewer non-smokers in the VA dataset compared to MIMIC.

**Table 2.** Distribution of task labels in the validation datasets.

| Task | Label | Primary validation, N=500 (VA) | External validation, N=500 (MIMIC) |
| --- | --- | --- | --- |
| Status | Current | 236 (47.2%) | 106 (21.2%) |
|  | Former | 121 (24.2%) | 188 (37.6%) |
|  | Non-Smoker | 34 (6.8%) | 153 (30.6%) |
|  | N/A | 109 (21.8%) | 53 (10.6%) |
| Duration ≥ 20 years | TRUE | 72 (14.4%) | 55 (11%) |
|  | FALSE | 428 (85.6%) | 445 (89%) |
| Quit period < 15 years | TRUE | 60 (12%) | 63 (12.6%) |
|  | FALSE | 440 (88%) | 437 (87.4%) |
| Intensity (pack per day) | <1 | 35 (7%) | 22 (4.4%) |
|  | 1 | 50 (10%) | 29 (5.8%) |
|  | 1–2 | 21 (4.2%) | 24 (4.8%) |
|  | >2 | 7 (1.4%) | 20 (4%) |
|  | N/A | 387 (77.4%) | 405 (81%) |
| Pack-years ≥ 20 | TRUE | 35 (7%) | 53 (10.6%) |
|  | FALSE | 465 (93%) | 447 (89.4%) |

**Table 3.** Overall patient cohort and dataset demographics.

|  | VA total |  | Dataset |  |  |  |  |  |
| --- | --- | --- | --- | --- | --- | --- | --- | --- |
|  |  |  | Primary validation |  | Training |  | Clinical application |  |
|  | N | (%) | N | (%) | N | (%) | N | (%) |
| **Total No. of Patients** | 58803 |  | 500 |  | 80 |  | 2500 |  |
| **Demographics** |  |  |  |  |  |  |  |  |
| Age at last note (years) |  |  |  |  |  |  |  |  |
| Mean (SD) | 70.8 | 7.91 | 70.1 | 7.74 | 71.5 | 8.41 | 70.7 | 7.82 |
| Age strata |  |  |  |  |  |  |  |  |
| 50-80 years old | 52039 | 88.5 | 451 | 90.2 | 68 | 85.0 | 2216 | 88.6 |
| Not 50-80 years old | 6764 | 11.5 | 49 | 9.8 | 12 | 15.0 | 284 | 11.4 |
| Sex |  |  |  |  |  |  |  |  |
| Male | 56544 | 96.2 | 485 | 97.0 | 75 | 93.8 | 2419 | 96.8 |
| Female | 2259 | 3.8 | 15 | 3.0 | 5 | 6.2 | 81 | 3.2 |
| **Snippets per patient** |  |  | 1 |  | 1 |  | 8.12 |  |

|  | MIMIC total |  | Dataset |  |
| --- | --- | --- | --- | --- |
|  |  |  | External validation |  |
|  | N | (%) | N | (%) |
| **Total No. of Patients** | 23500 |  | 500 |  |
| **Demographics** |  |  |  |  |
| Age at last note (years) |  |  |  |  |
| Mean (SD) | 66.08 | 8.88 | 66.19 | 8.92 |
| Age strata |  |  |  |  |
| 50-80 years old | 23500 | 100 | 500 | 100 |
| Not 50-80 years old | 0 | 0 | 0 | 0 |
| Sex |  |  |  |  |
| Male | 13960 | 59.4 | 287 | 57.4 |
| Female | 9540 | 40.6 | 213 | 42.6 |
| **Snippets per patient** |  |  | 1 |  |

Our zero-shot prompt showed a range of performance across tasks (Figure 1a), with a median F1 difference of 0.20 (interquartile range [IQR]:0.13-0.30) between tasks in the VA dataset and 0.10 (IQR:0.07-0.16) in MIMIC (Figure 1c). Zero-shot performance still exceeded the 2023 THYME temporal reasoning benchmark^34,29^, which did not use modern LLMs (macro-F1=0.63 in VA, 0.69 in MIMIC, 0.54 for THYME). Performance also differed between VA and MIMIC, with a median F1 difference of 0.09 (IQR:0.08-0.14) (Figure 1d). Although the LLM generally performed better on MIMIC data, this advantage was not consistent across tasks, indicating dataset- and task-specific factors may contribute to variability in performance and thus limit the generalizability of zero-shot results.

**Figure 1.**
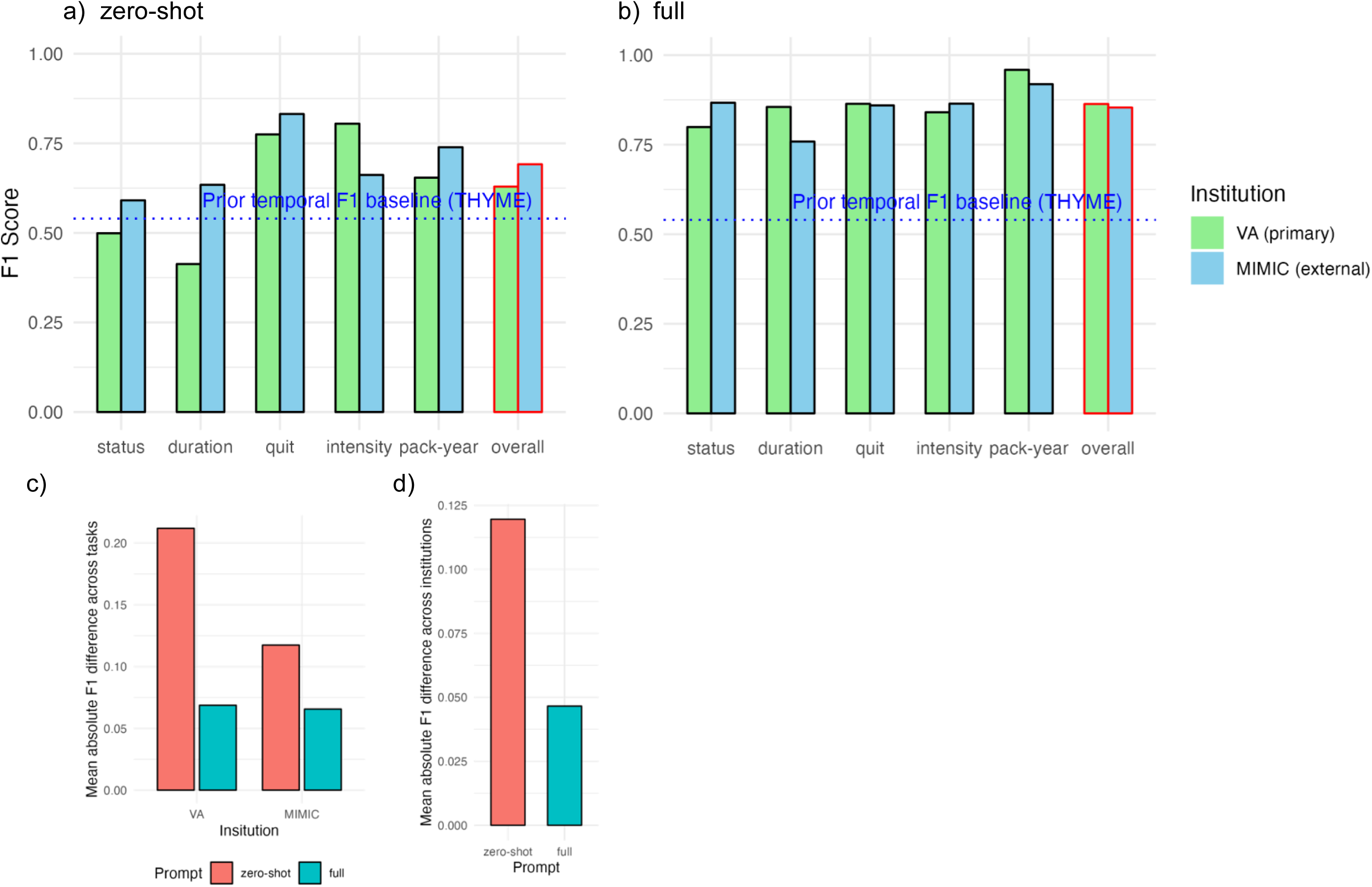
**Prompt engineering improves performance and generalizes across institutions** Performance of the a) zero-shot prompt and b) fully prompt engineered system on Mixtral 8×22B for five smoking tasks on the primary validation (VA, green) and external validation (MIMIC, blue) datasets. F1 scores are shown for binary outcomes (duration, quit year, pack-year), while macro-F1 scores (unweighted average of F1 scores across classes) are reported for multi-class outcomes (status, intensity). The “Overall” column represents the unweighted average F1 across all tasks (macro-F1, red outline). The dotted blue line represents the 2023 THYME2 temporal reasoning end-to- end NLP benchmark (F1=0.54). Also shown are the mean absolute F1 score differences: c) between tasks for the same institution and (d) between institutions for the same tasks, averaged across tasks. These differences are compared between the fully prompt engineered system (teal) and zero-shot (pink).

### Prompt engineering improves performance on all tasks and generalizes across institutions

We evaluated the potential of prompt engineering to adapt the smoking NLP system for increased performance and generalizability. We iteratively tested and improved prompts using a set of 80 note snippets from the VA that was separate from the 500 validation snippets. The final smoking NLP system consisted of a three-step prompt chain for extracting smoking time intervals and a single dedicated prompt for extracting pack-years. The first two prompts identified the start, duration, and end of smoking intervals, with a smoking intensity for each interval. The third prompt integrated this information with external data, such as the note date or birth date. Pack-years were extracted independently using a single dedicated prompt, as it did not require the temporal reasoning and calculations handled by the prompt chain.

System performance was validated on the same notes used for zero-shot testing. Prompt engineering improved performance across tasks (Figure 1b). Despite being developed only on VA data, the system performed comparably on MIMIC notes (macro-F1=0.86 in VA, 0.85 in MIMIC, Figure 1b) and reduced inter-task variability within MIMIC (Figure 1c). Generalizability across institutions improved, with the median F1 difference between VA and MIMIC decreasing from 0.09 (IQR:0.08-0.14) (zero-shot) to 0.04 (IQR:0.02-0.07) (Figure 1d).

### Prompt engineering improves performance additively on simple tasks and synergistically on complex tasks

To evaluate the impact of specific prompt engineering techniques on performance, we conducted a prompt ablation study. We systematically removed distinct sections from each of the four prompts in the system, with sections categorized based on the primary prompt engineering technique applied: additional specific instructions to provide context (”context”)^35^, output of intermediate reasoning steps (”show-your-work”)^36^, and examples for the model to follow (”in-context-learning”)^37^ (Figure 2a, Supplemental Figure 1).

**Figure 2.**
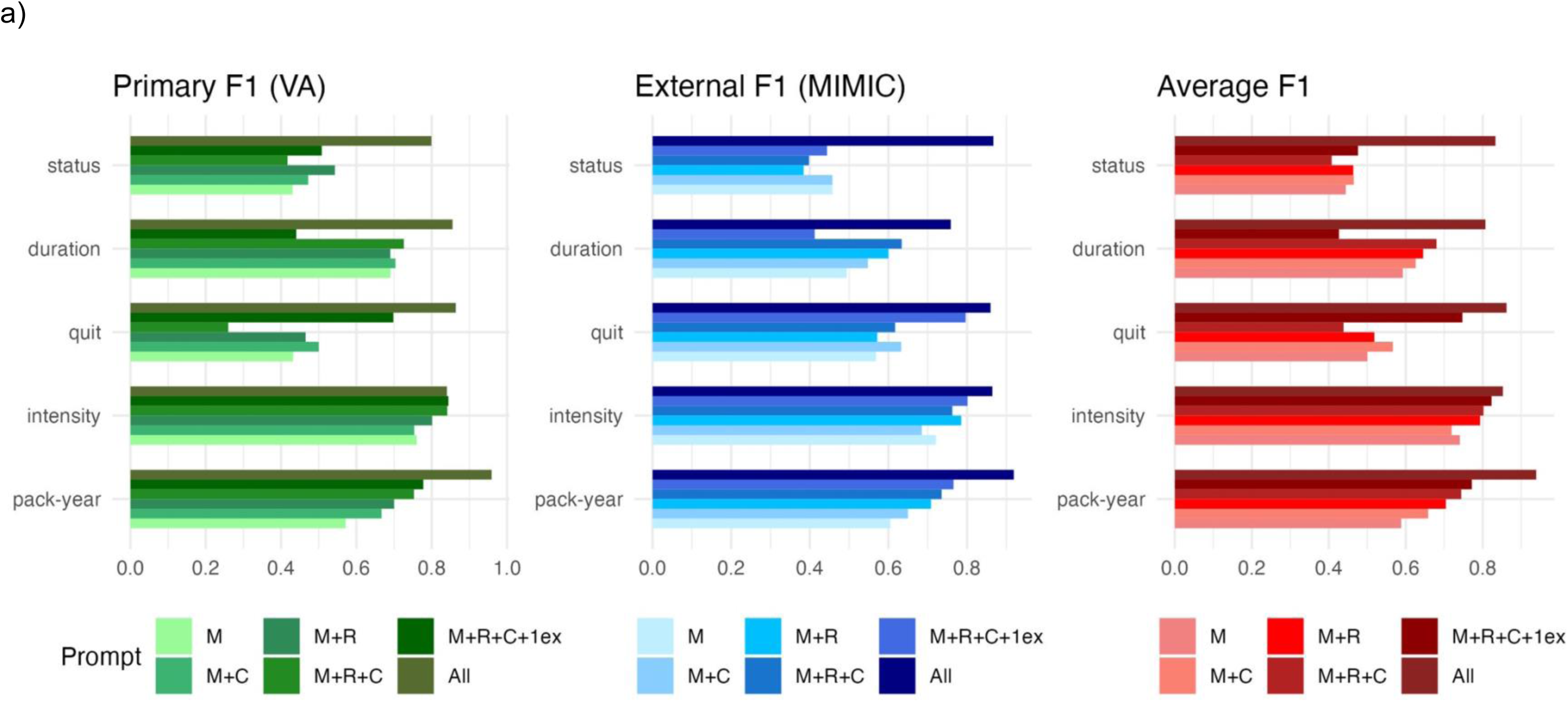
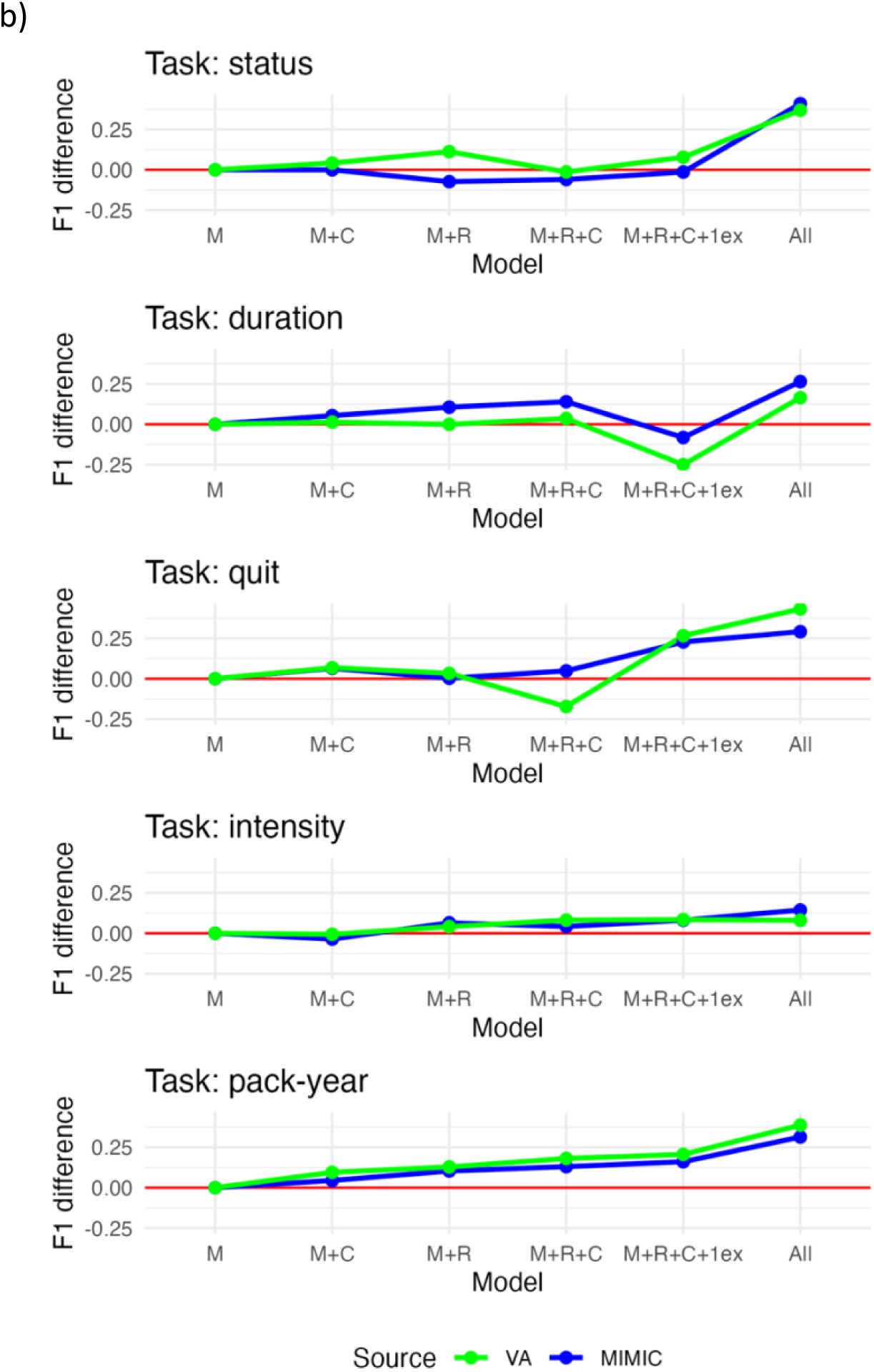
Prompt ablation study shows incremental improvements with each prompting technique on simpler tasks and synergistic improvement on complex tasks. Performance of the full prompt and prompt ablations on Mixtral 8×22B for five smoking tasks on the primary validation (VA, green) and external validation (MIMIC, blue) datasets. a) F1 scores are shown in for binary outcomes (duration, quit year, pack-year), while macro-F1 scores (unweighted average of F1 scores across classes) are shown for multi-class outcomes (status, intensity). The “Average F1” (red) represents the per-task unweighted average F1 across the institutions. B) The F1 score difference between each prompt and the minimal prompt (“F1 difference”) is graphed for each task. The solid red line represents a difference of zero. M = minimal prompt; C = additional context; R = intermediate reasoning steps; 1ex = one in-context learning example; All = full prompt with all components and additional in-context learning examples.

Within the single pack-years prompt, performance improved consistently with each added technique across datasets (Figure 2bc). In contrast, the three-prompt chain exhibited task-specific performance fluctuations with each added technique (Figure 2bc). Interestingly, intensity extraction, part of the chain, showed incremental improvements, unlike other tasks in the chain (Figure 2c). These findings suggest simpler tasks benefit linearly from added techniques, while complex, chained tasks may have synergistic gains from combined techniques.

### Hazards to LLM reasoning are unequally distributed across institutions

To identify dataset-specific hazards affecting LLM performance, we categorized errors from the final NLP system based on note input, intermediate reasoning, and final outputs. By analyzing intermediate reasoning outputs, we traced errors to distinct aspects of LLM behavior. Using qualitative content analysis, we developed a detailed codebook from observed errors and applied it to all 1,000 validation notes, including those without errors, to predict susceptibility to specific error types. This extended traditional error analysis by distinguishing errors from poor LLM performance on specific hazards versus high hazard prevalence in the dataset.

Our hazard analysis revealed significant differences in hazard distribution between the VA and MIMIC, despite comparable overall F1 scores (Figure 3a, Supplemental Table 1). Templated and minimally modified templated information were the most imbalanced hazards, with 132 instances in VA data compared to 27 in MIMIC (p_adj_=0.004 for both hazards). Moreover, snippets requiring advanced temporal reasoning for interpretation also imbalanced (p_adj_=0.008) and were more frequent in the VA dataset (17.5% of informative snippets) compared to MIMIC (4.6%) (Supplemental Figure 2).

**Figure 3.**
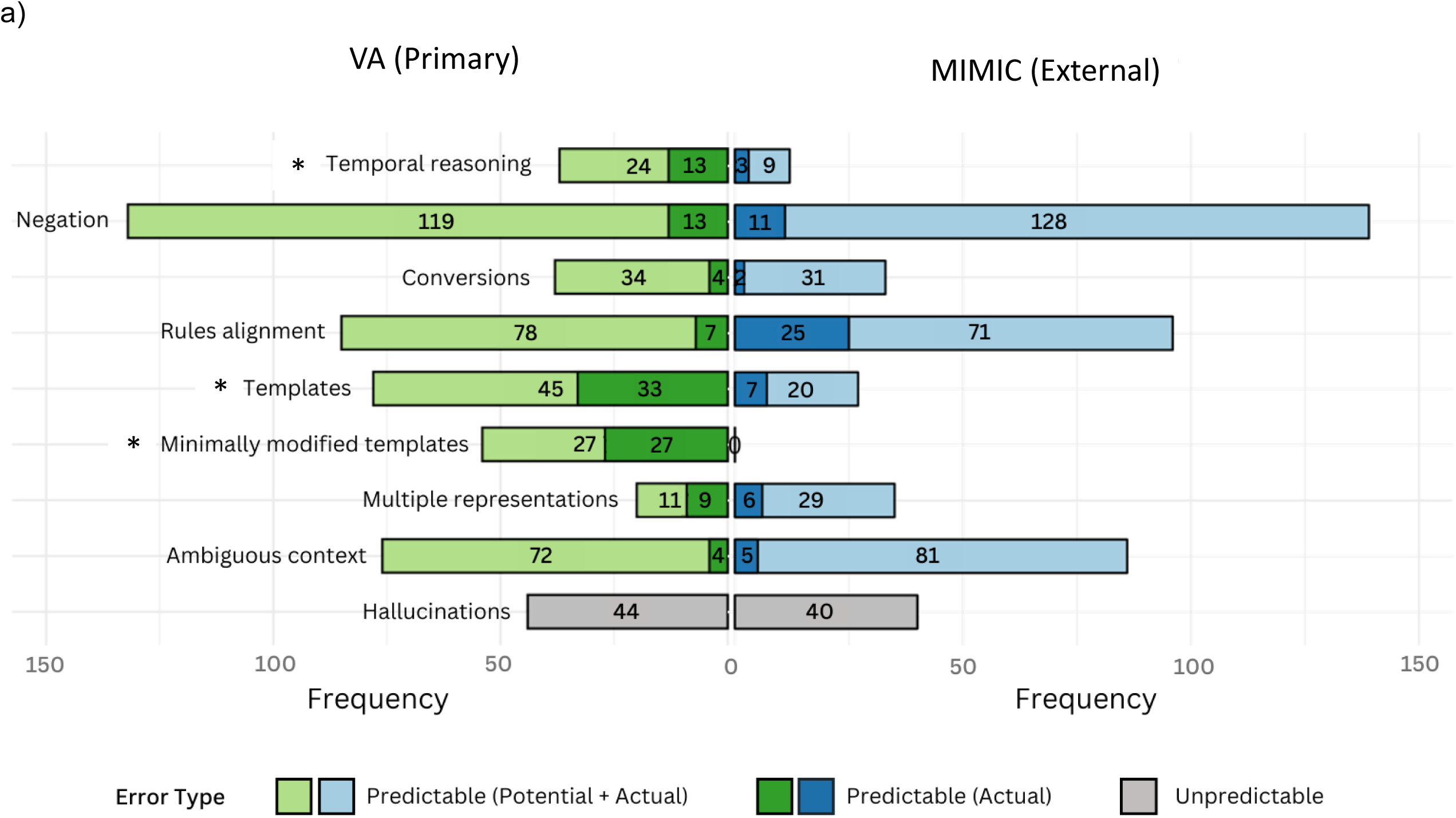
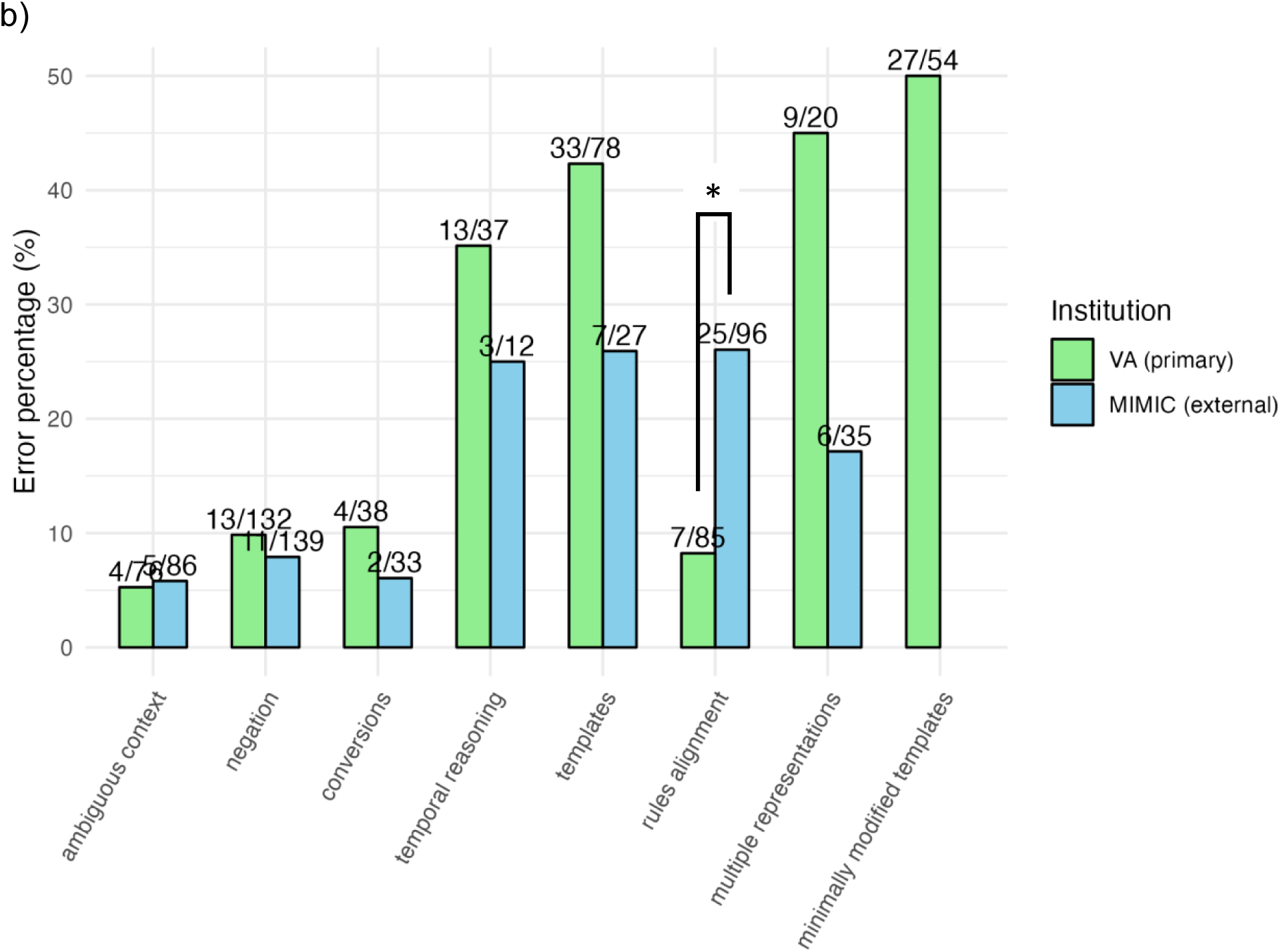
Hazards to LLM reasoning are unequally distributed across institutions. Hazard distribution (a) and error percentage by hazard (b) across validation datasets. Hazards were identified through qualitative content analysis of observed errors from the final NLP system, forming a comprehensive codebook applied to all validation notes, regardless of error presence. (a) Displays the overall distribution of hazards in the primary validation dataset (VA, green) and external validation dataset (MIMIC, blue). * represents p_adj_<0.05 for difference in overall hazard distribution between institutions by Chi-square test. (b) Shows the percentage of observed performance evaluation errors within each hazard category, separated by institution (VA and MIMIC). Arranged in increasing order of difference between error percentages by institution, except for minimally modified templates, which were not present in the MIMIC dataset. * represents p_adj_<0.05 for the difference in error percentages between institutions by two-proportion Z-test.

We calculated the percentage of errors for each hazard category to assess performance across datasets (Figure 3b, Supplemental Table 2). Overall, the system performed similarly or better on MIMIC data compared to VA data, despite being optimized exclusively on VA notes, with most hazards showing non-significant differences in error percentage. This suggests prompts targeting specific hazards can generalize across institutions. A notable exception was the “rules alignment” hazard (p_adj_=0.007), where the LLM generated plausible outputs that were incorrect under our annotation guidelines. The system performed better on VA data for this hazard, aligning with expectations from traditional training. These results indicate that the system generalizes well except in cases where annotation rules conflict with the LLM’s pre-trained behavior.

### Smoking history data extraction can identify patients who may be candidates for lung cancer screening

As a potential clinical application, we evaluated the utility of our NLP system to identify eligible candidates for lung cancer screening. Using standard VA data pipelines that are currently operational for national identification of eligible candidates, we could identify eligibility in only 8% of a 2500-person cohort of Veterans who later developed lung cancer. Our NLP algorithm increased eligibility identification of Veterans to 59% (1480/2500), 58% (861/1480) of whom were eligible for lung cancer screening under current USPSTF guidelines.

## Discussion

In this study, we identify and evaluate hazards to LLM generalizability between public and protected patient data and demonstrate how hazard-aware prompt engineering can mitigate them. We also address the unmet need for an end-to-end NLP system capable of extracting complex timelines from clinical text^29,34^. Notably, complex timelines are found in 17.5% of informative VA notes—nearly four times that of the most popular public dataset^7,25,27^, highlighting potential gaps in prior analyses. Our prompt-engineered system deploys multiple techniques to address LLM hazards like timelines, enabling such complicated tasks. Using this system, we demonstrate a 6.4-fold increase in lung cancer screening eligibility identification compared to current national VA tools.

Our findings demonstrate the feasibility of using prompt engineering to adapt LLMs for generalization across institutions, without fine-tuning or large training datasets. Our iterative prompt engineering process mirrored the hazard definition process, combining inductive (observing error patterns) and deductive (applying knowledge of clinical notes and LLM behavior) reasoning, formalized through our hazard codebook. Adaptation required only 80 notes—fewer than the hundreds or thousands typically needed with traditional methods^38^. Despite the small training size, the system generalized across institutions (VA and academic), time periods (2008–2021 for VA, 2001–2008 for MIMIC), and across 5 tasks and over 100 VA hospital systems, underscoring the potential of LLMs to build robust systems with minimal adaptation requirements.

Although our adapted prompt generalized well across institutions, the zero-shot prompt showed greater variation across tasks and datasets, appearing more vulnerable to imbalanced hazards between protected and public clinical data. For instance, LLM performance was poor on the hazard of templated information, such as social history templates, common in outpatient notes and VA data but less frequent in hospitalization-focused datasets like MIMIC^7,17^. This raises concerns that public data-based, general LLM benchmarks may not reflect realistic clinical scenarios or task-specific requirements^39^, potentially misaligning LLM development with clinical applications. Notably, our prompt engineering generalized worst when task-specific requirements conflicted with pre-trained model behavior, underscoring there is still a need to train or tune LLMs using real clinical needs and relevant patient data.

While further research could extend our approach to other tasks, smoking history serves as an illustrative example due to its long history with multiple comparators^22–27^, use of temporal reasoning central to clinical narratives^19,34^, and importance for preventing lung cancer morbidity and mortality through screening, which has less than 10% uptake^21^. A key barrier is the lack of accessible smoking data needed to determine eligibility, prompting the VA and other organizations to require manual entry into specialized EHR forms^40^. This process is time-intensive, error-prone, and duplicative, contributing to inefficiencies and burnout. As of September 2024, only 47% of current potentially eligible VA patients have documented smoking information^40,41^. Our LLM-based system addresses these gaps, outperforming manual processes and increasing coverage to 59%, potentially identifying screening eligibility in over a million additional Veterans.

Our study is not without limitations. By prioritizing detailed analysis of model behavior at each step of the reasoning pathway, the scope of clinical data extraction tasks is narrower. This focus is balanced by the diversity of notes analyzed^42^, the demonstrated range of task complexity, and the inclusion of nationally representative data. Additionally, our evaluation is limited to an open-source LLM. However, our zero-shot performance aligns with recent oncology benchmarks using GPT-4, indicating comparable capabilities with closed-source models^43^. While rapid model evolution may address some identified hazards, we defined hazards to reflect longstanding challenges in machine learning^34,44^ and thus remain relevant for understanding pathologies in smaller, more economical models needed for large-scale deployment.

In conclusion, LLMs show potential to generalize across public and protected patient data, though dataset-specific hazards can impact performance. Prompt engineering can address these hazards, as demonstrated by our smoking history extraction test case, with potential to automate lung cancer screening eligibility identification in a national healthcare system. As use of LLMs spreads across healthcare systems, defining and measuring the impact of these hazards provides a foundation for future efforts to improve performance, scalability, and deployment.

## Methods

### Patient and note selection and dataset generation

Our primary cohort consisted of Veterans diagnosed with lung cancer between March 1, 2018 and March 1, 2021. Our external validation MIMIC-III cohort consisted of 23,500 patients aged 50 to 80. We selected notes from both cohorts using established smoking-related keywords from prior smoking NLP studies^26^. Notes were included if they contained any smoking keyword, regardless of author or note type. For VA notes, we restricted selection to those recorded between 10 years and 6 months prior to lung cancer diagnosis. All MIMIC notes were included without timing restrictions. From the selected notes, we generated non-overlapping 200-character snippets centered around each smoking keyword.

From these snippets we generated the following distinct datasets:

1. **Primary validation:** 500 snippets, each from a different VA patient, with one randomly selected snippet per patient. Patients were selected to ensure an equal representation across VA healthcare systems.
2. **External validation:** 500 snippets, each from a different MIMIC patient, with one randomly selected snippet per patient
3. **Training:** 80 snippets, each from a different VA patient, distinct from the primary validation set, with one randomly selected snippet per patient
4. **Clinical application:** All snippets containing a smoking keyword from 2500 VA patients, including multiple snippets per patient when applicable. Patients were selected to ensure an equal representation across VA healthcare systems.

### Annotation of validation datasets

To establish a gold standard for model evaluation, we defined smoking intervals by start, duration, intensity, and end (Supplemental Figure 3). Smoking history tasks were defined from annotated smoking intervals as follows:

1. **Status**: current, former, or non-smoker. If the note mentions no current use of tobacco without a clear indication of smoking in the past, that patient was labeled a non-smoker.
2. **Duration**: length of smoking in years, calculated from the interval’s “start” and “end” (e.g., “smoked from age 14 to 2019”) or coded directly if explicitly stated (e.g., “smoked for 30 years”).
3. **Intensity**: number of packs smoked per day.
4. **Quit year**: time since quitting, coded directly (e.g., “quit for 15 years”) or calculated using external information as needed (e.g., “quit since 2019”)
5. **Pack-years**: pack-year history is defined as packs per day multiplied by years of smoking. We annotated only if explicitly stated in the snippet (e.g., “50 pk-yrs”) and excluded if calculations were required (e.g., “smoked 1pp for 50 years”) to minimize propagated errors from inferred values.

### Prompt engineering to develop the NLP system

For creation of the initial prompt, we structured a zero-shot prompt modeled after prompts published to perform well with note data on ChatGPT^32,33^ (Supplemental Figure 4). This zero-shot prompt included role play, clear section delimitation, and use of an example to specify desired output format.

For iterative prompt engineering, we tested the initial prompt on the 80 training dataset snippets and manually analyzed its performance, identifying common themes in misclassifications. Errors were addressed through iterative revisions until performance stabilized. Observing that combining multiple tasks in a single prompt limited performance—likely in part due to context window constraints—we split the tasks into a prompt chain for interval extraction and a separate prompt for pack-years.

### NLP performance evaluation

We evaluated the overall performance of initial prompt, final prompt chain system, and prompt ablations against our annotations on the primary validation (VA) and external validation (MIMIC) datasets. Snippets labeled with multiple intervals were manually reviewed to determine whether the NLP output matched annotations. For this performance evaluation, task outputs were categorized using thresholds based on the United States Preventative Services Task Force (USPSTF) guidelines:

1. **Status:** current, former, non-smoker
2. **Duration:** ≥20 years (TRUE or FALSE)
3. **Quit period:** <15 years (TRUE or FALSE)
4. **Intensity:** <1, 1, 1–2, >2 packs per day
5. **Pack-years:** ≥20 pack-years (TRUE or FALSE)

### Error analysis and hazard analysis

#### Error analysis

We conducted a qualitative analysis of exact-match errors by comparing task outputs from our annotations to those of the final NLP system on the primary (VA, N=500) and external (MIMIC, N=500) validation datasets. For each observed error, we compared the snippet, the NLP final output, and the four intermediate outputs from each prompt against the original annotation (total notes and LLM outputs = 1,955; Supplemental Figure 5). Errors identified during evaluation of the VA and MIMIC datasets across all five smoking tasks were analyzed using a combined inductive and deductive content analysis approach^45^ to generate a comprehensive codebook of LLM hazards based on observed errors.

#### Hazard annotation and analysis

We applied the codebook to identify inherent dataset hazards, focusing on instances with a predictable potential for incorrect smoking information extraction. Hazards were defined as predictable if the snippet content and codebook indicated an identifiable opportunity for error. Unpredictable errors, where no snippet information suggested a potential for error, were defined as “hallucinations.” Not all coding-phase errors were annotated as hazards, as some exact-match errors fell within the same cutoff-based bin used for NLP performance (F1 score) evaluation.

The primary (N=500) and external (N=500) validation datasets were analyzed using the codebook to identify hazards. The qualitative researcher who annotated the smoking tasks and developed the hazard codebook conducted the analysis, with ambiguous cases resolved by the lead researcher. Snippets could be assigned multiple hazards, and observed errors were automatically linked to their associated hazards. We used the Chi-square test to compare hazard distributions across datasets. Error percentages per hazard were evaluated using the two-proportion Z-test. Multiple hypothesis testing was adjusted for with the Bonferroni correction.

## Data Availability

All data produced in the present study are available upon reasonable request to the authors.

## Acknowledgements

We thank Travis Osterman for the fruitful correspondence by answering questions we had regarding their prior work on smoking history NLP and providing code. We thank VA Boston data scientists Kyle McGrath and Cenk Yildrim for assistance in setting up the large language model programming interface and executing data queries. They were not compensated.

